# Seroprevalence of dengue, Zika, yellow fever and West Nile viruses in Senegal, West Africa

**DOI:** 10.1101/2024.11.21.24317738

**Authors:** Sebastian Gallon, Mouhamad Sy, Prince Baffour Tonto, Ibrahima Mbaye Ndiaye, Mariama Toure, Amy Gaye, Mariama Aidara, Amadou Moctar Mbaye, Abdoulaye Kane Dia, Mamadou Alpha Diallo, Jules Francois Gomis, Mamadou Samba Yade, Younous Diedhiou, Baba Dieye, Khadim Diongue, Mame Cheikh Seck, Aida S. Badiane, Daouda Ndiaye, Bobby Brooke Herrera

## Abstract

West Africa serves as a critical region for the co-circulation of mosquito-borne flaviviruses, which often precipitate sporadic outbreaks. This study investigated the seroprevalence of dengue virus serotypes 1-4 (DENV-1-4), Zika virus (ZIKV), yellow fever virus (YFV), and West Nile virus (WNV) in three regions of Senegal: Sindia, Thies, and Kedougou. We retrospectively analyzed 470 serum samples for flavivirus immunoglobulin G (IgG) using a DENV-2 envelope (E) ELISA. Our findings revealed a seroprevalence of 37.23% for DENV-2 E IgG, indicative of a prior flavivirus exposure rate. The IgG seroprevalence rates for DENV-1-4, ZIKV, YFV, or WNV NS1 were 57.14%, 12.57%, 80.57% and 17.14%, respectively, with 72% of individuals harboring neutralizing antibodies against two or more flaviviruses. We also identified that residing in Sindia (ZIKV, OR, 9.428; 95% CI: 1.882-47.223 & WNV, OR, 6.039; 95% CI: 1.855-19.658) and Kedougou (ZIKV, OR, 7.487; 95% CI: 1.658-33.808 & WNV, OR, 1.142; 95% CI: 0.412-3.164) was a significant risk factor for ZIKV and WNV exposure. In contrast, history of malaria significantly reduced the risk of WNV exposure (aOR, 0.402; 95% CI: 0.203-0.794). This study underscores the complexity of flavivirus epidemiology in West Africa and the necessity for enhanced surveillance to inform public health strategies.

## Introduction

Flavivirus, a genus within the *Flaviviridae* family, comprises over 70 arthropod-borne viruses characterized by a single-stranded, positive-sense RNA genome.^1^ Notable members of this group include dengue virus serotypes 1-4 (DENV-1-4), Zika virus (ZIKV), yellow fever virus (YFV), and West Nile virus (WNV), all of which pose significant global health risks. Transmission occurs via distinct mosquito vectors, with *Aedes* species primarily responsible for spreading DENV-1-4^2^, ZIKV^3^ and YFV^4^, while *Culex* mosquitoes primarily transmit WNV.^5^ The Flavivirus genome encodes for a single polyprotein, composed of three structural (capsid, envelope [E], and pre-membrane) and seven non-structural (NS1, NS2A, NS2B, NS3, NS4A, NS4B, and NS5) proteins.^6^ Phylogenetic analysis suggests that all vector-borne flaviviruses originated in Africa, where mosquito vectors likely played a central role in their evolution and spread.^7^ The prevalence of these mosquitoes is further intensified by warm, humid climates and human-driven environmental changes, such as deforestation, which create ideal breeding conditions that can facilitate their spread into rapidly urbanizing areas.^8^ In many resource-constrained regions, limited healthcare infrastructure hampers the diagnosis and treatment of flavivirus infections.^8^ Moreover, the clinical symptoms of flavivirus infections often overlap with those of other endemic diseases, like malaria or typhoid, leading to underreporting or misdiagnosis in the absence of specific acute-phase symptoms.^9^

In Africa, where DENV-1-4 is endemic in at least 34 countries^10^, infections are frequently inapparent or manifest as mild disease.^11^ However, outside of Africa, dengue is responsible for significant morbidity and mortality, with millions of annual cases characterized by a spectrum of clinical manifestations, ranging from mild dengue fever to severe forms such as dengue hemorrhagic fever and dengue shock syndrome.^12^ Despite its global prevalence, the true burden of dengue in Africa remains poorly understood due to inadequate diagnostic and surveillance infrastructure.^13^

In Senegal, the first documented dengue case occurred in 1970, underscoring its emerging public health importance.^14^ All four dengue serotypes maintain sylvatic transmission cycles and are associated with sporadic outbreaks.^15^ Historically, DENV-2 has predominated, but the epidemiological landscape shifted in 2009, when DENV-3 emerged, particularly in urban areas like Thies.^16^ In recent years, the situation has escalated to hyperendemic levels, with annual outbreaks driven by multiple serotypes, such as the 2017-2018 outbreak of DENV-1-3.^17^ The availability of three polyvalent dengue vaccines signals progress in controlling dengue outbreaks, but only the Sanofi Pasteur dengue vaccine has been approved for use in Africa.^18^ However, Sanofi Pasteur’s recent announcement to halt manufacturing of the dengue vaccine emphasizes the urgent need for robust surveillance to mitigate future outbreaks across Africa.

YFV, believed to have originated in African rainforests, has plagued human populations for centuries and was first isolated during the 1927 epidemic in West Africa.^19^ This strain served as the basis for the development of the YF17D vaccine, a live-attenuated vaccine that induces long-lasting immunity in 99% of vaccinated individuals.^20^ In countries such as Senegal, vaccination campaigns targeting early childhood have achieved coverage rates of 80-100% in specific regions.^21^ However, inconsistencies in vaccination coverage persist, leading to periodic outbreaks, such as those that occurred in eastern Senegal between 2020-2021.^22^ These gaps in coverage are attributed to limited healthcare infrastructure, suboptimal vaccine distribution, and under-resourced immunization campaigns.^21^ Consequently, YFV remains a major public health threat in West Africa, particularly in unvaccinated populations, with an estimated annual burden of 84,000 to 170,000 severe cases.^23^ Surveillance of populations for neutralizing antibodies, whether from vaccination or prior infection, is crucial for assessing seroprevalence and understanding the role of pre-existing immunity in modulating responses to subsequent flavivirus infections.

ZIKV, first identified in Uganda in 1947, has been studied intermittently in Senegal from 1962 through 2018.^24, 25^ As a neurotropic virus, ZIKV is linked to severe outcomes, such as microcephaly in neonates^26^ and Guillain Barre syndrome in adults.^27^ The 2015-2016 Latin American ZIKV outbreak highlighted the importance of tracking emerging neglected tropical diseases. Initially, only Asian lineage ZIKV strains were thought to be associated with neurological complications in infants, but recent studies indicate that African lineage ZIKV may exhibit greater transmissibility and pathogenicity.^28^ A subsequent study found that ZIKV infections in pregnant women in Nigeria were associated with adverse neonatal outcomes, including microcephaly and other congenital anomalies.^26^ In Senegal, ZIKV primarily circulates within a sylvatic transmission cycle involving non-human primates and arboreal *Aedes* mosquitoes, with human cases predominantly occurring in rural areas.^29^ Despite this, there have been no large-scale human outbreaks reported in Africa, and seroprevalence studies in Senegal have documented relatively low ZIKV seropositivity rates, ranging from 7.5%-13%, suggesting low-level transmission.^30^

WNV, also first identified in Uganda in 1947, circulates primarily in an enzootic cycle between birds and mosquitoes.^31^ With nine genetically distinct lineages identified, WNV exhibits considerable genetic diversity, and lineages 1, 2, 7, and 8 have been reported circulating in Africa.^31^ Although most WNV infections are inapparent or mild, a subset of cases results in severe neurological manifestations, including encephalitis.^31^ Notably, a large WNV outbreak affected thousands of individuals in South Africa in 1974, and sporadic outbreaks continue to occur across Africa.^32^ While WNV has been documented in Senegal, no major outbreaks have been reported, despite serological evidence of its circulation. Additional in depth seroprevalence studies are needed to assess the current burden of WNV in Senegal.

Given the increasing threat posed by flavivirus infections across Africa, there is a need to prioritize surveillance efforts and to better characterize the immune responses elicited by these infections. The objective of this retrospective study was to determine the seroprevalence of DENV-1-4, ZIKV, YFV, and WNV in non-febrile serum collected from individuals residing in three regions in Senegal. Furthermore, we assessed the neutralizing capacity of each serum against these viruses. Our findings suggest a high frequency of individuals experiencing multiple and/or sequential flavivirus infections, potentially harboring cross-neutralizing antibodies, which has significant implications for public health and vaccine development strategies.

## Materials and Methods

### Study Populations and Ethics Statement

The samples included in our study were originally collected as part of malaria and non-malarial surveillance among residents of Sindia, Thies, and Kedougou, Senegal. Samples from Sindia and Thies were part of the surveillance of non-malarial febrile illnesses, while samples from Kedougou were part of genomic surveillance of malaria. Informed consent was obtained from all subjects and/or their legal guardians for the initial sample collection as well as for its future use. All methods were performed in accordance with the guidelines and regulations set forth by the Declaration of Helsinki.

The primary studies under which the samples and data were collected received ethical clearance from the CIGASS Institutional Review Board (IRB) (Protocol numbers: SEN15/46, 19300, and SEN14/49). All excess samples and corresponding data were banked and de-identified prior to the analyses. This study received and exemption determination from the Rutgers Robert Wood Johnson Medical School IRB.

### ELISAs

Human sera were subjected to DENV-2 E Immunoglobulin G (IgG) enzyme-linked immunosorbent assay (ELISA, as previously described^25^) to test for the presence of DENV-2 E IgG antibodies to determine flavivirus serostatus. Nunc Maxisorp 96 well plates (Thermofisher) were coated with DENV-2 E (20 ng per well; Native Antigen) overnight. Subsequently, plates were blocked for 1 hour with 1x PBST, 1% BSA (Thermofisher) and then washed with 1x PBST a total of three times. Plates were then incubated with primary antibodies for 2 hours at room temperature (serum diluted 1:400). Plates were subject to washing three times to wash unbound antibody and bound sera IgGs reacted with secondary antibodies (anti-human IgG conjugated with horseradish peroxidase [HRP]; Thermofisher), according to the manufacturer’s instructions. The plates were washing three more times with 1X PBST, then allowed to settle with 1-step TMB ELISA substrate (Thermofisher), per the manufacturer’s instructions. An ELISA reader (Diasource) with a test OD of 450 nm and a reference wavelength of OD 650 nm was used to interpret the ELISA.

Subsequently, NS1-based IgG ELISAs (as previously described in^25^) were performed to determine the seropositivity rates against DENV1-4, ZIKV, YFV and WNV among the DENV-2 E IgG+ serum samples. Similarly to DENV-2 E-based ELISA, 96 well plates were individually coated with DENV1-4, ZIKV, YFV, or WNV NS1 proteins (20 ng per well; Native Antigen), blocked, washed, and treated with primary antibodies (serum diluted 1:400). Bound sera IgGs reacted with secondary antibodies (anti-human IgG conjugates with HRP; Thermofisher) and plates were subsequently washed and treated with 1-step TMB ELISA substrate (Thermofisher). Plates were read with an OD of 450 nm and a reference wavelength of 650 nm.

### Microneutralization Assays

Microneutralization assays were used to determine the proportion of samples harboring neutralizing antibodies to a specific flavivirus. Serum samples were diluted in a two-fold series from 1:10 to 1:1280 in 1X PBS in a 96-well plate. For each dilution, contact was performed with an equivalent volume of a 100 plaque forming unit (PFU) dose of DENV-1 (Hawaii strain), DENV-2 (DakArA1247 strain), DENV-3 (CH53489 strain), DENV-4 (H241 strain), ZIKV (DAK AR 41524 strain), YFV (YFV17D strain), or WNV (Bird114 strain) for 90 minutes at 37°C in a 5% CO_2_ incubator. After neutralization, the complexes were then added to pre-seeded Vero cell monolayers (CCL-81, ATCC) in 96-well plates and an additional 100 μl of cell culture medium was added without removal of the virus inoculum after adsorption. After 3-6 (ZIKV, YFV, WNV) to 7-10 (DENV-1-4) days of incubation, the cytopathogenic effects were investigated. The cell culture medium was removed from the plates and the cells were stained with 0.2% crystal violet (SigmaAldrich) in 20% Ethanol (SigmaAldrich). After staining for 2 hours, the pates were washed with copious amounts of water.

The 90% neutralization titer (NT90) of the test serum sample against each virus was defined as the reciprocal of the test serum dilution for which the virus infectivity was reduced by 90% relative to the challenge virus dose (without any antibodies). A titer lower than 1:20 was considered as negative if cytopathogenic effects were not observed. For monotypic infections, samples were considered positive if the titer was ≥1:20. For samples that had neutralizing activity against two or more viruses, samples were considered positive if the titer was ≥1:40; a higher NT90 cutoff was used as a more stringent means for determination. Microneutralization of each virus was validated using immune serum from previously infected humans (BEI Resources) or mice.

Additionally, at least 3 flavivirus-naïve serum samples were used to incubate with DENV-1, DENV-2, DENV-3, DENV-4, ZIKV, YFV, or WNV prior to pipetting onto the Vero cells per plate, and 100 PFU of DENV-1, DENV-2, DENV-3, DENV-4, ZIKV, YFV, or WNV alone without any contact with serum were placed on Vero cells in triplicate per plate.

### Statistical Analysis

To investigate the association between flavivirus seroprevalence and demographic factors, such as age group, gender, geographic location, and malaria status, both univariate and multivariate binary logistic regression analyses were employed. These models allowed for the estimation of the odds ratios (ORs) for each independent variable, quantifying the likelihood of flavivirus seropositivity. Adjusted odds ratios (aORs) were calculated to account for potential confounders, specifically age and gender, ensuring that the observed associations were not biased by these variables.

Categorical variables, including seroprevalence and demographic characteristics, were summarized as case counts and percentages, while continuous variables, such as age, were presented as medians with accompanying ranges. The multivariate analysis included all covariates that were statistically significant in the univariate analysis or that were deemed important based on prior literature. Statistical significance was assessed at a p-value threshold of <0.05. All analyses were conducted using IBM SPSS Statistics software version 28, and 95% confidence intervals (CIs) were calculated for all odds ratios to assess the precision of the estimates.

To evaluate the performance of the NS1-based ELISAs, we compared their results to those obtained from the microneutralization assay, which served as the gold standard for determining true positive and true negative rates. Sensitivity and specificity were calculated based on the comparison of ELISA results with the microneutralization assay, with sensitivity representing the proportion of true positives correctly identified by the ELISA, and specificity representing the proportion of true negatives. The 95% confidence intervals for sensitivity and specificity were calculated using the exact Clopper-Pearson method to provide robust estimates of precision. Moreover, the heatmap and Venn diagram, representing neutralization titers against the flaviviruses, were generated by GraphPad Prism version 10.2.0 and Microsoft PowerPoint, respectively.

## Results

A total of 470 serum samples were collected from three regions in Senegal: Sindia, Thies, and Kedougou. Of these, 94 samples each were obtained from Sindia and Thies in 2018, while 282 samples were collected from Kedougou between 2022 and 2023. Participants’ median ages were 24 (range: 2-70 years) in Sindia, 20 (range: 6 months-69 years) in Thies, and 21 (range: 2-85 years) in Kedougou (Table 1). Samples from Sindia and Thies were from malaria-negative individuals, while all samples from Kedougou were from malaria-positive individuals (Table 1).

**Table 1.** Description of population characteristics and serology results of DENV-2 envelope (E) ELISA testing.

| Characteristics and Serology | Sindia<br>(n=94) | Thies<br>(n=94) | Kedougou<br>(n=282) | Total<br>(N=470) |
| --- | --- | --- | --- | --- |
| **Demographics** |  |  |  |  |
| Year(s) samples collected | 2018 | 2018 | 2022-2023 |  |
| Median age (range) | 24 (2-70) | 20 (6 months-69) | 21 (2-85) | 21 (6 months-85) |
| % Female (number) | 41.49 (39) | 54.25 (51) | 42.91 (121) | 44.89 (211) |
| **Malaria status, number (%)** |  |  |  |  |
| Positive | 0 | 0 | 282 (100) | 282 (60) |
| Negative | 94 (100) | 94 (100) | 0 | 188 (40) |
| **Flavivirus status (based on**<br>**DENV-2 E IgG), number (%)** |  |  |  |  |
| Positive | 33 (35.11) | 27 (28.72) | 115 (40.78) | 175 (37.23) |
| Negative | 61 (64.89) | 67 (71.28) | 167 (59.22) | 295 (62.77) |

Out of the total cohort, 175 samples (37.23%) positive for DENV-2 E IgG by ELISA (Table 1). Flavivirus seroprevalence was further differentiated using NS1-based IgG ELISAs for DENV-1-4, ZIKV, YFV, or WNV. In Sindia (n=33), the seroprevalence rates for DENV-1-4, ZIKV, YFV, and WNV were 75.76%, 9.09%, 69.70%, and 39.39%, respectively (Table 2). In Thies (n=27), DENV-1-4 seroprevalence was 62.96%, with ZIKV, YFV, and WNV seropositivity rates of 3.70%, 70.37%, and 14.81%, respectively (Table 2). In Kedougou (n=115), seroprevalence rates were 50.43% for DENV-1-4, 15.65% for ZIKV, 86.09% for YFV, and 11.30% for WNV (Table 2). Across all regions, pooled NS1 seroprevalence rates were calculated as 57.14% for DENV-1-4, 12.57% for ZIKV, 80.57% for YFV, and 17.14% for WNV (Table 2).

**Table 2.** Serology results of nonstructural protein 1 (NS1) ELISA testing.

|  | Sindia<br>n=33 | Thies<br>n=27 | Kedougou<br>n=115 | Total<br>N=175 |
| --- | --- | --- | --- | --- |
| DENV-1-4 NS1, n (%) | 25 (75.76) | 17 (62.96) | 58 (50.43) | 100 (57.14) |
| ZIKV NS1, n (%) | 3 (9.09) | 1 (3.70) | 18 (15.65) | 22 (12.57) |
| WNV NS1, n (%) | 13 (39.39) | 4 (14.81) | 13 (11.30) | 30 (17.14) |
| YFV NS1, n (%) | 23 (69.70) | 19 (70.37) | 99 (86.09) | 141 (80.57) |

To assess the reliability of the NS1-based ELISAs, we compared their performance against the neutralization assay. Sensitivity and specificity rates for each virus were determined. For pan-DENV, the sensitivity and specificity were 89.72% and 94.12%, respectively. For ZIKV, the sensitivity was 87.50%, and the specificity was 99.34%. For YFV, the sensitivity was 91.39%, with a specificity of 87.50%. For WNV, the sensitivity was 88.46% and the specificity was 95.30% (Supplementary Table 1). These results underscore the robust accuracy of NS1-based ELISAs in detecting flavivirus seropositivity.

Neutralization assays were performed to evaluate the presence of neutralizing antibodies (nAbs) across sample populations. The overall prevalence rates for nAbs against DENV-1, DENV-2, DENV-3, and DENV-4 were 18.85%, 12.57%, 32.57%, and 5.71%, respectively (Table 3). The combined prevalence of nAbs against any DENV serotype was 69.71% (Table 3). ZIKV, YFV, and WNV nAb prevalence rates were 13.71%, 86.28%, and 14.85%, respectively (Table 3). In Sindia, nAb rates against DENV-1, DENV-2, DENV-3, and DENV-4 were 30.03%, 15.15%, 15.15%, and 12.12%, respectively, with an overall pan-DENV nAb rate of 72.72% (Table 3). ZIKV, YFV, and WNV nAb rates were 12.12%, 93.93%, and 36.36%, respectively (Table 3). In Thies, the nAb rates against DENV-1, DENV-2, DENV-3, and DENV-4 were 18.51%, 22.22%, 37.03%, and 0%, respectively, with a pan-DENV nAb rate of 77.77% (Table 3). ZIKV, YFV, and WNV nAb rates were 3.70%, 66.66%, and 11.11%, respectively (Table 3). In Kedougou, nAb rates against DENV-1, DENV-2, DENV-3, and DENV-4 were 15.65%, 9.56%, 36.52%, and 5.21%, respectively, with a pan-DENV nAb rate of 66.95% (Table 3). ZIKV, YFV, and WNV neutralization rates were 16.52%, 88.69%, and 9.56%, respectively (Table 3).

**Table 3.** Neutralizing capacity to DENV-1-4, ZIKV, YFV, and WNV by study site determined by NT90 titers.

|  | **Sindia**<br>**(n=33)** | **Thies**<br>**(n=27)** | **Kedougou**<br>**(n=115)** | **Total**<br>**(N=175)** |
| --- | --- | --- | --- | --- |
| **DENV-1, n (%)** | 10 (30.03) | 5 (18.51) | 18 (15.65) | 33 (18.85) |
| **DENV-2, n (%)** | 5 (15.15) | 6 (22.22) | 11 (9.56) | 22 (12.57) |
| **DENV-3, n (%)** | 5 (15.15) | 10 (37.03) | 42 (36.52) | 57 (32.57) |
| **DENV-4, n (%)** | 4 (12.12) | 0 (0) | 6 (5.21) | 10 (5.71) |
| **pan-DENV,**<br>**n (%)** | 24 (72.72) | 21 (77.77) | 77 (66.95) | 122 (69.71) |
| **ZIKV, n (%)** | 4 (12.12) | 1 (3.7) | 19 (16.52) | 24 (13.71) |
| **WNV, n (%)** | 12 (36.36) | 3 (11.11) | 11 (9.56) | 26 (14.85) |
| **YFV, n (%)** | 31 (93.93) | 18 (66.66) | 102 (88.69) | 151 (86.28) |
| **Negative, n (%)** | 2 (6.06) | 4 (14.81) | 7 (6.08) | 13 (7.42) |

The NT90 data revealed a high prevalence of nAbs, with 72% of subjects displaying neutralizing capacity against two or more flaviviruses (Fig. 1A). In particular, 13.14% of subjects produced nAbs against three or more flaviviruses. NAbs to DENV and YFV were observed in 32.0% of cases, while 6.85% of subjects exhibited neutralization patterns corresponding to secondary DENV (sDENV) plus YFV (Fig. 1B). Additionally, 4.57% of subjects demonstrated neutralization against ZIKV and YFV, while 7.42% had nAbs against both WNV and YFV.

**Figure 1.**
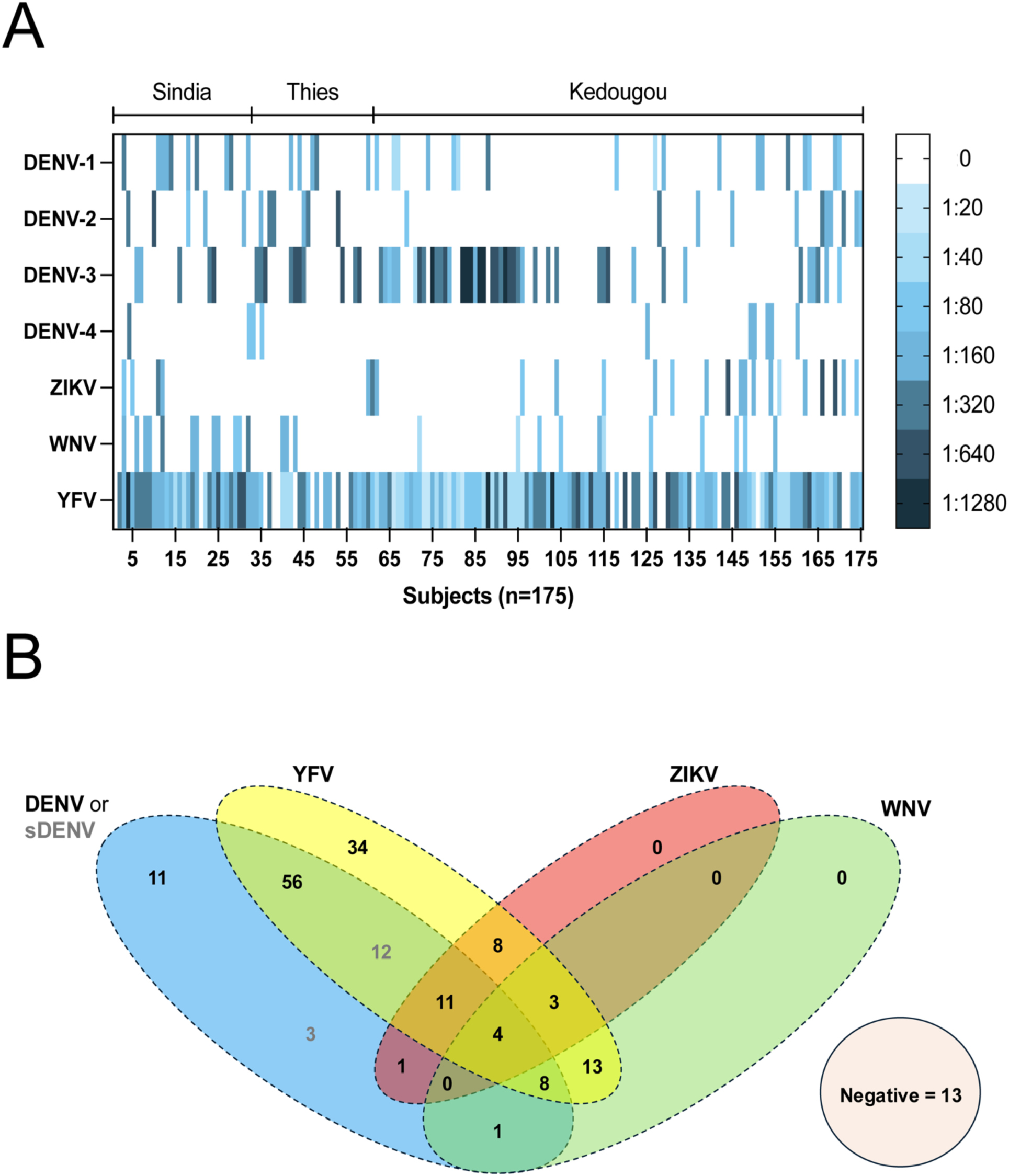
**A.** Neutralizing titers across all three cohorts. Abbreviations: DENV (dengue virus), sDENV (secondary dengue virus), YFV (yellow fever virus), ZIKV (Zika virus) and WNV (West Nile virus). The representative sample size is 175 subjects whose serum was tested for neutralizing antibodies to all flaviviruses included in this study. **B.** Flavivirus specific neutralization rates from positive PRNT90 results. 13 samples were negative for neutralization against all viruses tested.

Multivariate logistic regression analysis identified several factors associated with flavivirus infection. Individuals aged 21–40 years (OR, 2.830; 95% CI: 1.855–4.318) and those over 40 years old (OR, 3.379; 95% CI: 1.797–6.353) were more likely to have flavivirus exposure based on DENV-2 E IgG reactivity, with the highest risk in individuals over 40. Geographic location also played a role; residents of Sindia (OR, 1.323; 95% CI: 0.691–2.534) and Kedougou (OR, 1.782; 95% CI: 1.034–3.072) had increased odds of flavivirus exposure. Furthermore, a history of malaria significantly increased the likelihood of flavivirus infection (aOR, 3.465; 95% CI: 2.253–5.328) (Table 4).

**Table 4:**
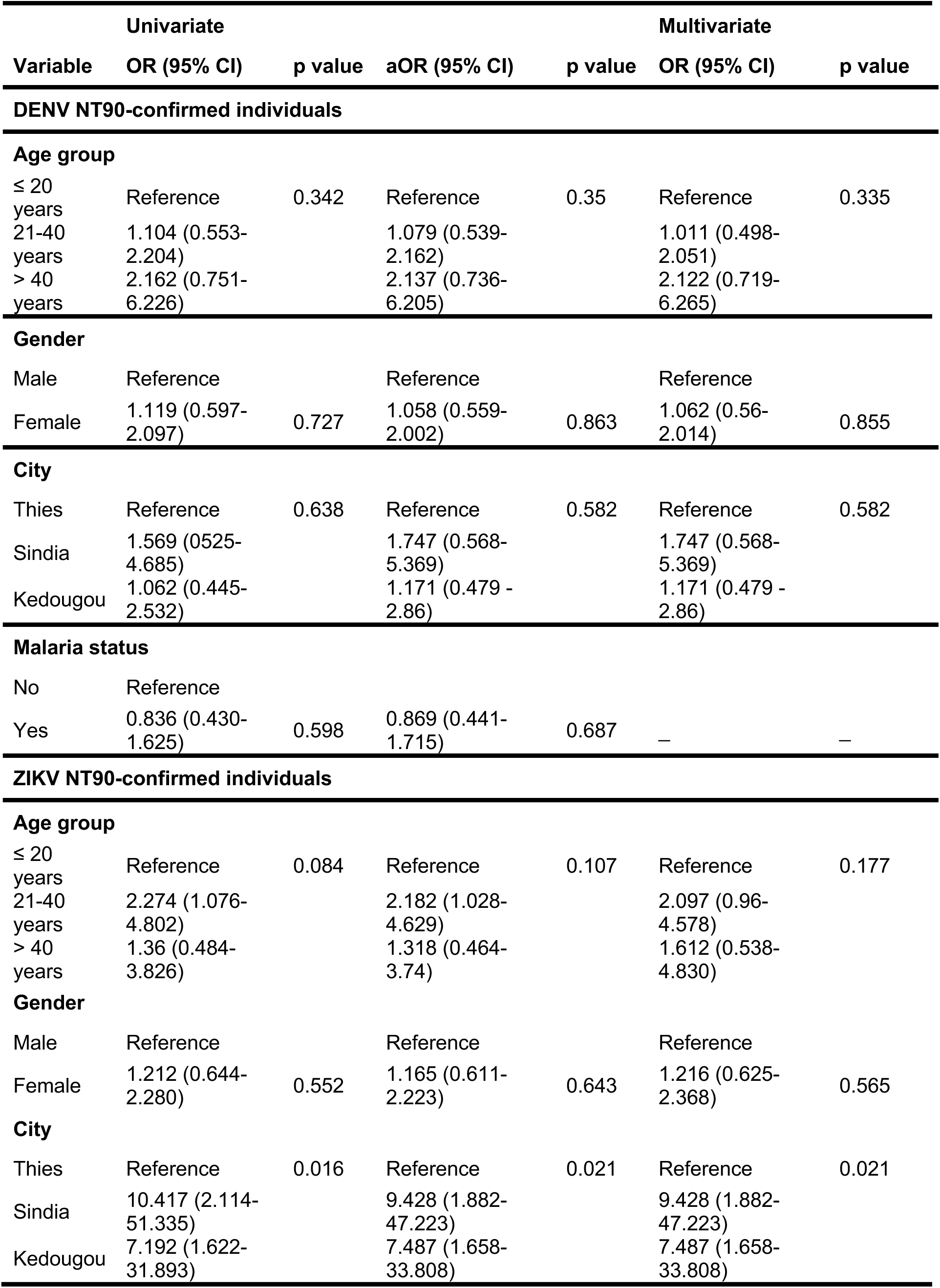
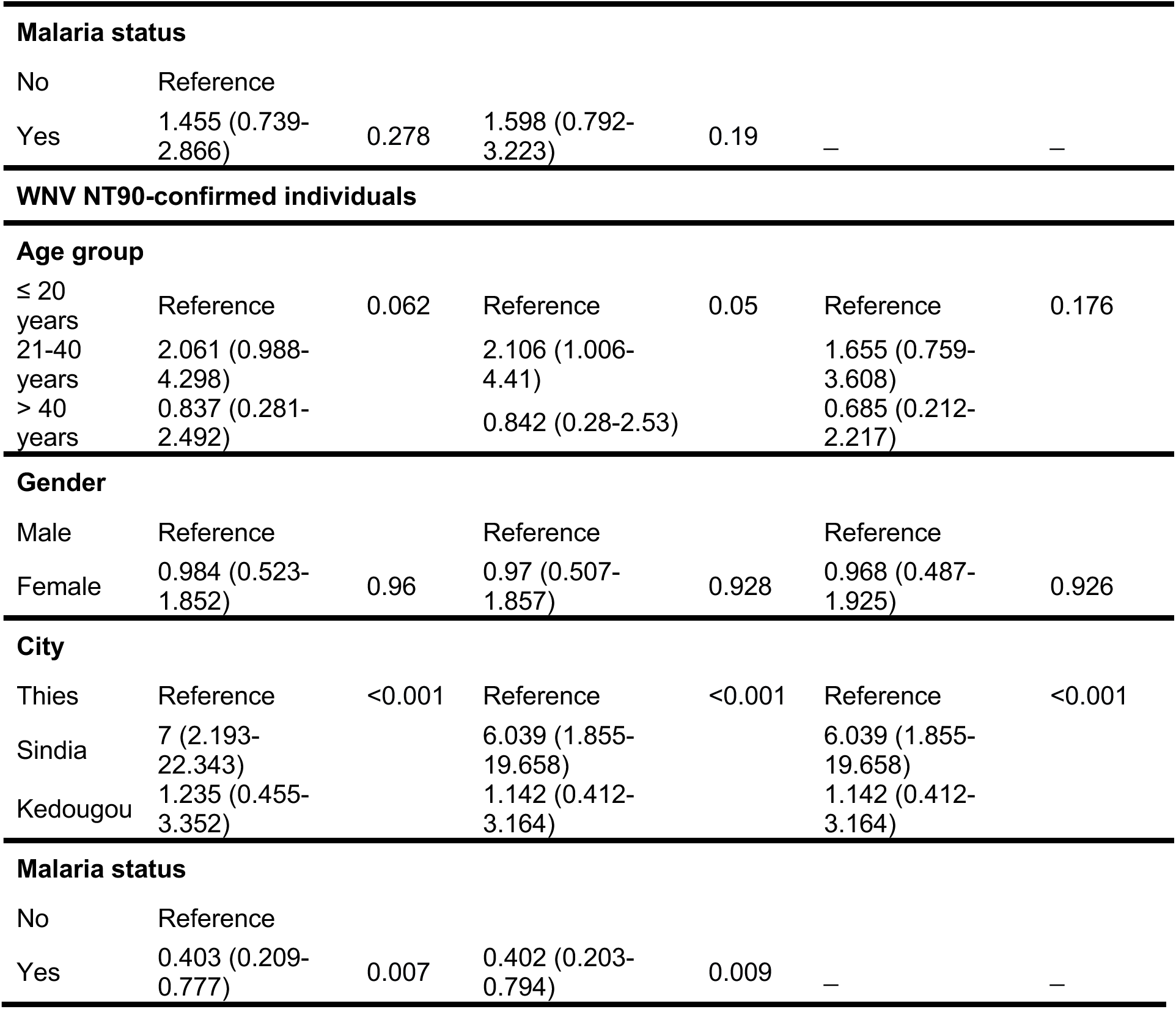
Risk factors for DENV-1-4, ZIKV and WNV exposure in all study sites.

In contrast, logistic regression models indicated no significant association between DENV infection risk and other modeled variables. However, Sindia residents had a significantly higher risk of ZIKV infection (OR, 9.428; 95% CI: 1.882–47.223), while residents of Kedougou had a heightened risk of WNV infection (OR, 7.487; 95% CI: 1.658–33.808), with lower odds of WNV infection in individuals with a history of malaria (aOR, 0.402; 95% CI: 0.203–0.794) (Supplementary Table 2).

## Discussion

This study on the seroprevalence of DENV-1-4, ZIKV, YFV, and WNV in Senegal highlights the heterogenous distribution of these flaviviruses, driven by geographical and ecological factors that influence transmission dynamics. Among the 470 serum samples analyzed, 37.23% exhibited seropositivity to DENV-2 E, suggestive of prior flavivirus exposure, with the highest seroprevalence noted in Kedougou. This aligns with the region’s longstanding status as an endemic zone, where extensive sylvatic surveillance since the 1960s has contributed valuable insights into the evolutionary ecology and circulation of arboviruses in these complex ecosystems.^33^ Understanding these dynamics is crucial for the development of targeted public health interventions that reflect the transmission nuances of difference regions.

The high degree of conservation within the flavivirus E protein, exhibiting up to 40% sequence homology across species, poses significant challenges for serological differentiation.^34^ While E-based ELISAs are commonly used due to their cost-effectiveness and ability to detect broad flavivirus exposure, their specificity is limited by cross-reactivity – a drawback exacerbated in regions where multiple flaviviruses co-circulate.^37, 38^ Nevertheless, our DENV-2 E-based ELISA provided a robust estimate of overall flavivirus seroprevalence (37.23%) within the study population, confirming prior flaviviral exposure rates in Sindia (35.11%), Thies (28.72%), and Kedougou (40.78%) (Table 1).

The implementation of NS1-based IgG ELISAs, which target the less conserved non-structural protein, has served as a valuable tool for distinguishing between virus-specific infections.^37–39^ In this study, NS1-based ELISAs demonstrated high sensitivity and specificity (Supplementary Table 1), corroborated by neutralization assays that remain the gold standard for confirming prior exposure.^40^ The results obtained underscore the necessity of employing complementary methods, such as neutralization assays, to confirm specific flavivirus infections and minimize diagnostic ambiguities.

Our findings revealed significant exposure to YFV, with NS1 seroprevalence rates of 86.09% in Kedougou, compared to 69.70% and 70.37% in Sindia and Thies, respectively (Table 2). Overall, the YFV nAb rate was 86.28%, supporting high YFV exposure, either through YFV17D vaccination or natural infection (Table 3, Fig. 1). This is consistent with prior reports documenting multiple distinct YFV lineages in Senegal, particularly concentrated in Kedougou, a recognized YFV hotspot due to its expansive forest habitats facilitating sylvatic transmission.^41^ Also, the Senegalese government’s vaccination campaigns, including the 2021 mass vaccination initiative, likely influenced these high seroprevalence rates.^22^ Such efforts are essential for understanding regional immunity patterns and the persistence of YFV across different landscapes.

The DENV-1-4 NS1 seroprevalence was notably high in Sindia (75.76%), Thies (62.96%), and Kedougou (50.43%), reflecting the broad endemicity of DENV across Senegal. Urban centers, such as Thies, experience intensified transmission due to *Aedes aegypti* proliferation, whereas sylvatic transmission also contributes to persistent DENV circulation with outbreak potential in areas like Kedougou.^42, 43^ The 2018 outbreak^44^, which heavily impacted Sindia and Thies, likely contributed to these elevated seroprevalence rates, a finding supported by the high prevalence of nAbs observed against DENV-1 (30.03%) in Sindia, and DENV-3 (37.03%) in Thies (Table 3, Fig. 1). Overall, the pan-DENV nAb rate was 69.71% across all subjects, with rates ranging from 5.71% for DENV-4 to 32.57% for DENV-3 (Table 3, Fig. 1).

ZIKV NS1 seroprevalence across all study sites was 12.57%, comparable to findings from similar studies in Senegal, which reported seropositivity rates of approximately 13%.^30^ The presence of ZIKV nAbs in 13.71% of subjects corroborates these findings (Table 3, Fig. 1). Moreover, WNV exposure was also evident, with NS1 seroprevalence rates of 14.81% in Thies and 11.30% in Kedougou, but markedly higher in Sindia (39.39%). Similarly, the presence of WNV nAbs in 14.85% of subjects supports these findings (Table 3, Fig. 1). Previous studies have demonstrated widespread WNV circulation among mosquito vectors and enzootic hosts, such as birds and horses, across Senegal.^45^ Although WNV outbreaks have not been extensively reported, the evidence of nAbs indicates sporadic human exposure and the potential for future outbreaks.^31^ Further studies need to be performed to assess other risk factors that may pre-dispose some locations in Senegal to higher WNV exposure.

Moreover, our neutralization data also provided insights into co-infections, sequential infections, and/or cross-neutralization. Most subjects (72%) exhibited neutralizing activity against two or more flaviviruses, and 2.28% of subjects demonstrated potential cross-neutralization against all tested flaviviruses. This finding underscores the complexity of immune responses in regions with persistent flavivirus co-circulation. Differentiating between secondary DENV infections and potential DENV-ZIKV co-infections remains challenging due to their substantial cross-reactivity. Notably, 13 subjects showed seropositivity in the DENV-2 E ELISA, but lacked nAbs, suggesting waning antibody levels that no longer confer neutralization, as reported in a prior study.^46^ Another possible expalanation that cannot be excluded is false positivity of the DENV-2 E-based ELISA. However, the potential waning of YFV nAbs post-vaccination, particularily in infants and young children, has also been observed^47^, warranting further investigation into the duration of immunity following natural infection versus YFV17D vaccination.

The role of pre-existing immunity in modulating subsequent flavivirus infections is of particular interest. Cross-reactivity between DENV and YFV, and its potential protective or detrimental effects, has been documented in multiple studies.^48, 49^ Our findings support this complex interplay, with individuals showing cross-neutralization between DENV and YFV potentially reflecting an adaptive immune response that influences subsequent exposure outcomes. The lack of monotypic ZIKV and WNV infections further emphasizes the possibility that pre-existing immunity to more prevalent flaviviruses, such as DENV, or through YFV17D immunization, may attenuate ZIKV or WNV outbreaks.

Our multivariate logistic regression analysis did not reveal any significant association between DENV exposure and demographic factors, such as age, gender, location, or malaria status. However, our analysis revealed significant associations between geographic location and exposure risk, with Sindia and Kedougou residents showing higher odds of ZIKV and WNV exposure, respectively (Table 4). The association between a history of malaria and reduced WNV exposure presents an intriguing hypothesis that malaria-induced immunity may provide cross-protection, a phenomenon observed in a study examining SARS-CoV-2 cross-reactivity in a malaria endemic region of Senegal.^50^ However, the mechanisms behind this relationship remain speculative and require further exploration.

Limitations of this study include its retrospective design and the absence of clinical data, preventing confirmation of recent or acute infections through IgM testing. Additionally, distinguishing YFV17D vaccination-induced and natural infection-induced antibodies was not feasible, which may affect interpretations of seroprevalence data. Despite these limitations, this study represents one of the most comprehensive evaluations of DENV-1-4, ZIKV, YFV, and WNV seroprevalence in Senegal to date, further corroborated by neutralization data. The findings emphasize the need for continued surveillance and deeper investigation into flavivirus molecular epidemiology, including flavivirus genome analyses, in West Africa, particularly in areas with overlapping viral circulation and vaccination campaigns.

In conclusion, this study sheds light on the complex interactions between flaviviruses in Senegal, shaped by ecological, historical, and immunological factors. The high rates of flavivirus neutralization observed in our study underscore the need for ongoing research into the immune responses elicited by concurrent flavivirus exposures. Understanding these interactions will be critical for developing more effective vaccines and public health strategies to manage outbreaks in flavivirus-endemic regions.

## Data Availability

Data produced as part of the current study are available in the manuscript or upon request from the corresponding author.

## Acknowledgements

We would like to thank Rutgers Global Health Institute, Rutgers Robert Wood Johnson Medical School, and The Child Health Institute of New Jersey for their continued support. The funders had no role in writing of the manuscript or decision to publish the manuscript.

## Competing Interests Statement

BBH is a co-founder of Mir Biosciences, Inc., a biotechnology company focused on T cell-based diagnostics and vaccines for infectious diseases, cancer, and autoimmunity.

## Supplementary Tables

**Supplementary Table 1.**
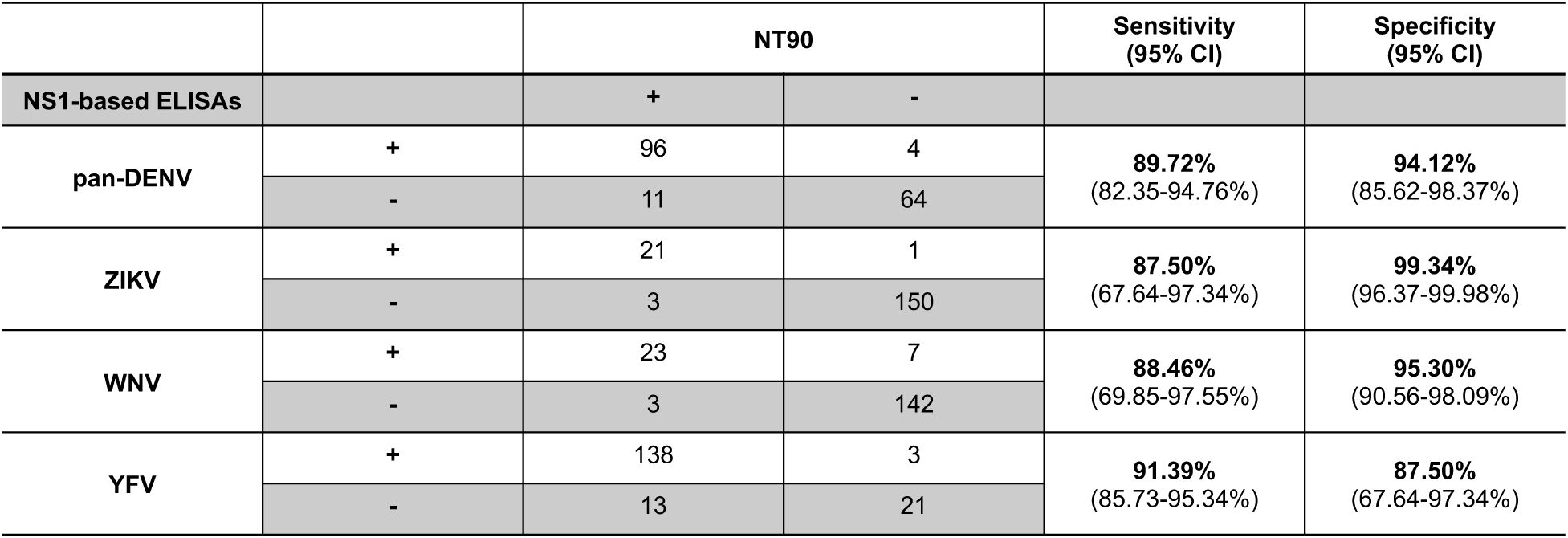
Sensitivity and specificity values calculated from NS1-based ELISAs and NT90 results.

**Supplementary Table 2.**
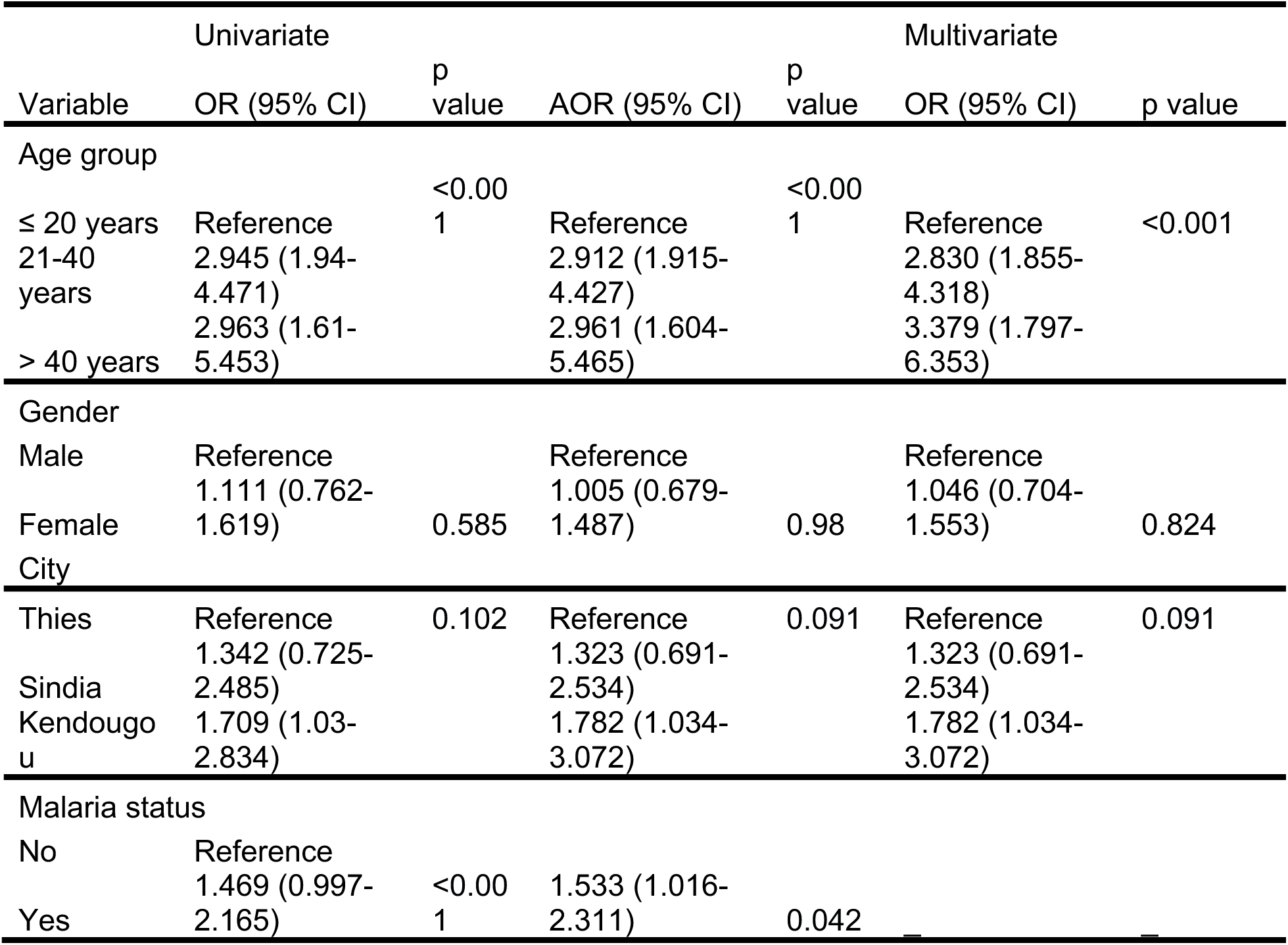
Univariate vs multivariate regression analysis.

## Notes

### Author Declarations

The samples included in our study were originally collected as part of malaria and non-malarial surveillance among residents of Sindia, Thies, and Kedougou, Senegal. Samples from Sindia and Thies were part of the surveillance of non-malarial febrile illnesses, while samples from Kedougou were part of genomic surveillance of malaria. Informed consent was obtained from all subjects and/or their legal guardians for the initial sample collection as well as for its future use. All methods were performed in accordance with the guidelines and regulations set forth by the Declaration of Helsinki. The primary studies under which the samples and data were collected received ethical clearance from the CIGASS Institutional Review Board (IRB) (Protocol numbers: SEN15/46, 19300, and SEN14/49). All excess samples and corresponding data were banked and de-identified prior to the analyses. This study received and exemption determination from the Rutgers Robert Wood Johnson Medical School IRB.

